# Seroprevalence of RSV Antibodies in a Contemporary (2022 – 2023) Cohort of Adults

**DOI:** 10.1101/2025.01.14.25320554

**Authors:** Lara I. Teodoro, Inna G. Ovsyannikova, Diane E. Grill, Gregory A. Poland, Richard B. Kennedy

**Author notes:** **Address correspondence to:** Richard B. Kennedy, PhD, 200 1st St SW, Rochester, MN 55905.

## Abstract

**Objectives:** Respiratory syncytial virus (RSV) infection may cause serious illness and mortality in older adults (≥ 65 years). This study aimed to assess the seroprevalence of anti-RSV IgG antibodies in adults from a United States cohort and compare antibody levels among individuals with a history of recent RSV vaccination or infection.

**Methods:** A total of 475 subjects (ages 27 – 99) were randomly selected from the Mayo Clinic Biobank’s residual sera repository (2022 – 2023). Samples were tested for anti-RSV IgG using an enzyme-linked immunosorbent assay (ELISA). Additional cohorts included individuals with documented RSV infection (n = 40) or recent RSV vaccination (n = 71).

**Results:** Among the seroprevalence cohort, 83.8% tested RSV IgG positive. Males had significantly higher antibody titers than females (p < 0.001), and antibody titers increased post-RSV season (p = 0.02). Compared to the general cohort, RSV-seropositivity rates were higher in recently diagnosed (97.5%) and vaccinated (95.8%) individuals.

**Conclusions:** This study demonstrates a high seroprevalence of RSV IgG in adults, with variations across sex and seasonality, and corroborates waning immunity following RSV infection. Recent infection or vaccination significantly boosts antibody levels, reinforcing the importance of continued surveillance of RSV immunity.

## 1. INTRODUCTION

Respiratory syncytial virus (RSV) pertains to the family *Pneumoviridae*, which contains negative-strand RNA viruses known for infecting the lungs and respiratory tract.[1] RSV has two subtypes, A and B, each with slightly different antigenic properties,[2] although a significant amount of antibody cross-reactivity has been found. The National Institute of Allergy and Infectious Diseases (NIAID) estimates that almost all children in the US will have been infected with RSV by two years of age.[3, 4] RSV infection causes cold-like symptoms in healthy individuals.[5] However, infection in very young children and in older adults (≥ 65 years old) may cause severe disease, complications, and death.[6] The Centers for Disease Control and Prevention (CDC) estimates that RSV causes over 60,000 hospitalizations and 6,000 deaths each year among US adults ≥ 65 years of age. The durability of protection after natural infection is low – intranasal RSV challenge studies demonstrated that 47% of adults could be reinfected as early as 2 months after natural infection.[7] Although neutralizing antibody titers ≥ 6.0 log_2_ are expected to prevent RSV-associated hospitalizations[8] and monoclonal antibodies are protective, a correlate of protection has not yet been established.

Three single-dose RSV vaccines have been approved by the US Food and Drug Administration (FDA) since 2023.[9] Each vaccine is based on the RSV F glycoprotein but utilizes distinct antigen sources and/or delivery technologies. Arexvy (GSK) consists of pre-fusion stabilized F glycoprotein from RSV A and contains the AS01E adjuvant.[10] Abrysvo (Pfizer) consists of recombinant pre-fusion stabilized F glycoproteins from both strain groups (RSV A and RSV B).[11] MRESVIA (Moderna) consists of nucleoside-modified mRNA encoding pre-fusion stabilized F glycoprotein from RSV A in a lipid nanoparticle.[12]

Serosurveillance studies estimate a population’s level of exposure to a pathogen and can consequently be used to optimize the deployment and use of vaccines in that population. Previous studies on the incidence of RSV infection among adults in the United States focused exclusively on individuals who sought medical care or were hospitalized for acute respiratory illness or related symptoms.[13-21] Our study sought to assess the seroprevalence of RSV antibodies in a broader population, using a cohort of adults with available serum samples, who were not selected based on the presence of acute respiratory illness or related symptoms. Therefore, it is complementary to existing data from other geographic regions and published studies in U.S. cohorts and can be used to better understand the prevalence and patterns of RSV immunity in various population groups. The study cohort of 475 subjects (ages 27 – 99) was screened for RSV antibodies. Additional cohorts of subjects with a previous RSV diagnosis (N = 40) or recent RSV vaccination (N = 71) were studied for comparison.

## 2. METHODS

### 2.1 Study Cohort

The seroprevalence study cohort consisted of 475 adults (ages 27 – 99) residing in Rochester, Minnesota, or the surrounding areas. These subjects were obtained by cluster random sampling of residual sera collected from Mayo Clinic Biobank participants (n = 9,006) who had a blood draw between January 2022 and November 2023 at Mayo Clinic. Samples were processed and stored at -80°C. All sera samples were included in the sample collection, regardless of the reason/cause for the blood draw. All study participants had provided written informed consent for the use of demographic and medical history (e.g., RSV vaccination and diagnosis records) data. The described procedures were approved by the Mayo Clinic Institutional Review Board (IRB# 08-007049). To create our cohort of 475 subjects, the biospecimens from the first 20 individuals obtained each month were selected under the assumption that the patients arrived in a random pattern for their blood draws. Exceptions to this were the use of 40 specimens from January 2022 (the initial month of the study) and June 2023 (to cover the lack of available samples from May 2023), as well as the exclusion of 5 samples from subjects who were later found to be repeats.

Additionally, the electronic health record of the 9,006 subjects from the residual sera repository was searched for a previous history of RSV diagnosis or RSV vaccination, yielding the additional cohorts of diagnosed (n = 40) and vaccinated (n = 71) subjects.

### 2.2 RSV ELISA

To assess serum anti-RSV IgG, an enzyme-linked immunosorbent assay (ELISA) coated with whole virus and viral components purified from Vero cells was performed using the human anti-respiratory syncytial virus IgG ELISA kit (AB108765, Abcam®, Sweden) following the manufacturer’s recommendations. Serum samples were prepared by diluting 1:1,000 in the sample diluent provided within the assay kit. Kit-supplied controls and standards were provided on the required concentrations and were plated without any additional dilution. Controls and standards were assayed in duplicate on each plate and used to calculate antibody titers in standard units, while each sample was also evaluated in duplicate.

Each well’s optical density (OD) was measured by a SpectraMax 6 plate reader at 450 nm (OD_450 nm_) and 620 nm (OD_620 nm_). The net OD for each well was determined by subtracting measured absorbance at OD_620 nm_ from OD_450nm_ (OD_450nm_ – OD_620 nm_). The average OD was calculated by averaging the net OD results of duplicates. Finally, standard units (SU) were calculated by multiplying the subject’s average OD by 10 and dividing the result by the average cut-off control’s OD [(subject’s average OD x 10) / average cut-off control OD]. The results were interpreted as negative (<9 SU), equivocal (9 – 11 SU), and positive (> 11 SU) as per the manufacturer’s recommendations. In our laboratory, the coefficient of variation for this assay was 2.4%.

### 2.3 Statistical Methods

Statistically significant differences between two groups were tested using Wilcoxon rank-sum tests. Kruskal-Wallis tests were used for comparisons involving more than two groups. Pearson’s correlation was used to calculate correlations between continuous variables. Analyses were conducted using the R statistical package version 4.4.1.

### 2.4 RSV Seasonality

The National Respiratory and Enteric Virus Surveillance System (NREVSS) dashboard was utilized to define the start and end of the RSV season in SE Minnesota for the years covered by our sample collection. The dataset included the percentage of RSV-positive test results weekly reported to NREVSS, which was filtered by pathogen (RSV) and by the US Department of Health and Human Services (HHS) Region (Region 5, encompassing Minnesota).[22, 23] The traditional 10% threshold was applied, defining the RSV season as the period when weekly positive percentages exceeded 10%.[24]

## 3. RESULTS

### 3.1 Study Cohort

The seroprevalence study cohort (N = 475) consisted mainly of white subjects (97.5%). The sex distribution was 183 (38.5%) males and 292 (61.5%) females. The age of participants at the time of blood collection ranged from 27 – 99 years, with a median of 72.8 (Q1: 61.9, Q3: 82.0) years. From this cohort, only one subject had a prior RSV diagnosis documented in their medical records, with the diagnosis occurring more than five years before sample collection. This subject was included in this cohort for the analysis. No subjects had records of RSV vaccination before sample collection. In addition to the main study cohort, subjects with a previous RSV diagnosis (N = 40; 55.0% female; median age 80.6) or recent vaccination record (N = 71; 59.2% female; median age 75.7 years) were tested. Table 1 presents detailed demographic information for the seroprevalence cohort, along with the diagnosed and vaccinated groups.

**Table 1.**
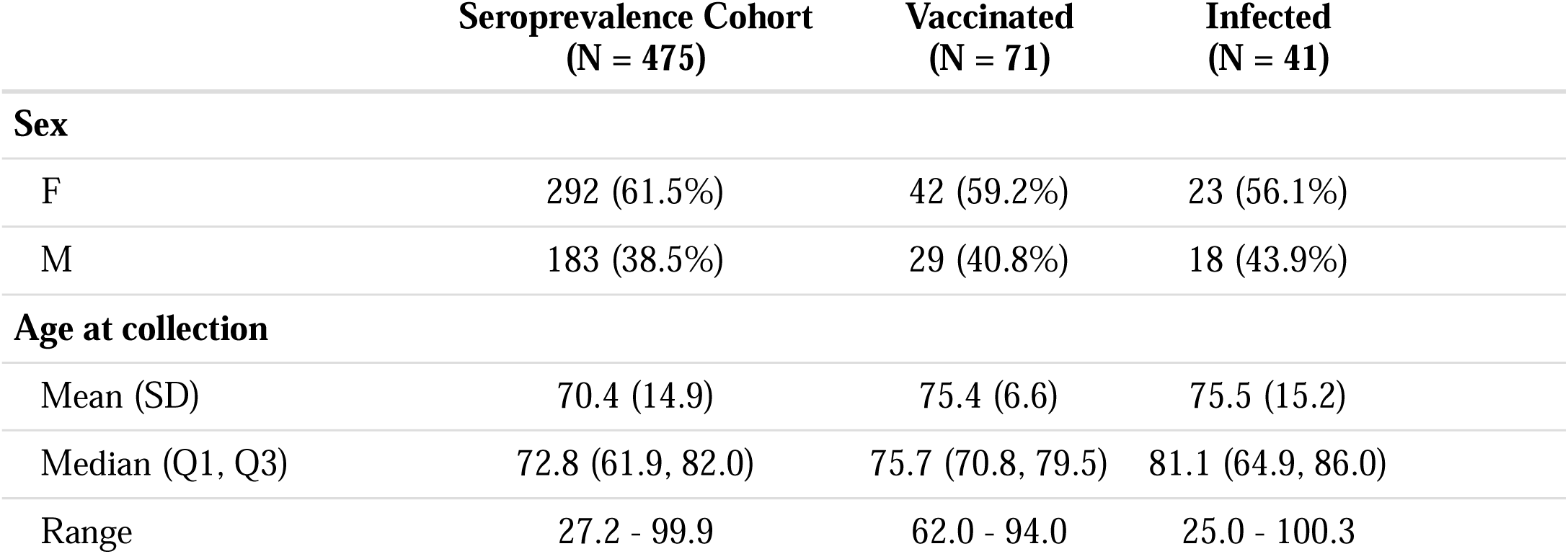
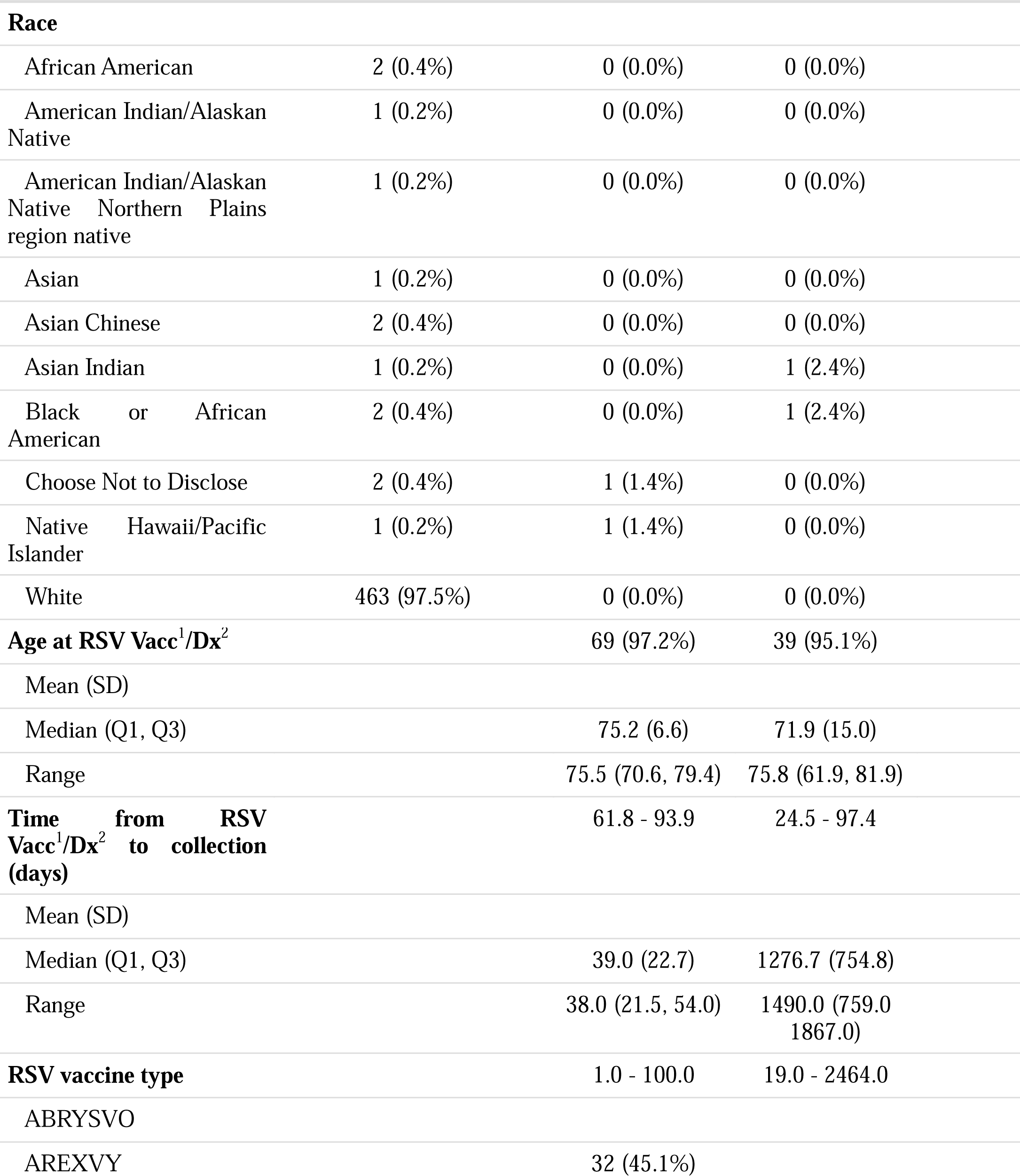
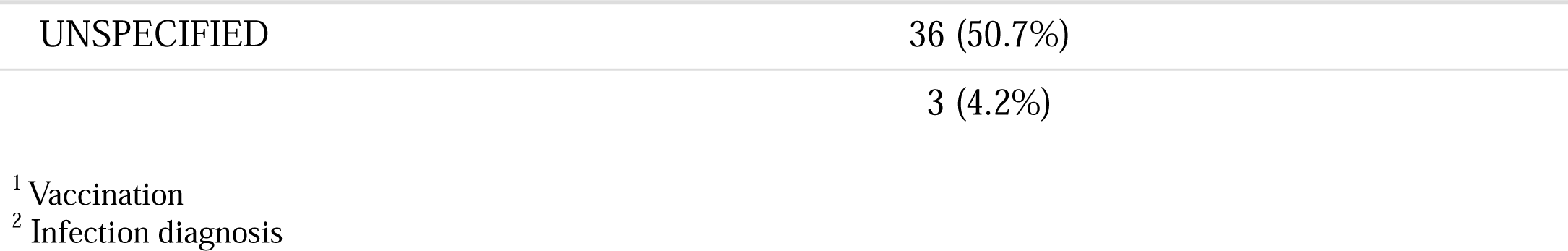
Demographics.

### 3.2 Seropositive Rates of RSV

Of the 475 subjects from the seroprevalence study cohort, 398 (83.8%) tested IgG positive, 38 (8.0%) tested IgG negative, and 39 (8.2%) had equivocal results for the presence of RSV IgG in their serum. Wilcoxon rank-sum test indicated that males had significantly higher median antibody titers than females (p < 0.001), with the median titer being 15.5 standard units (SU) for males and 14.5 SU for females. Figure 1 depicts the proportion of seroprevalence results by sex (Fig. 1A), the comparison of antibody titers by sex (Fig. 1B) or age group (Fig. 1C), and the comparison of antibody titers before, during, and after RSV season (Fig. 1D). There were no statistical differences between subjects with records of potential immunocompromising conditions and those without such records (SU min: 3.2 vs 4.0; SU max: 39.8 vs 40.8; median: 15.5 vs 15.8; p > 0.5) (Supplemental Figure 1).

**Figure 1.**
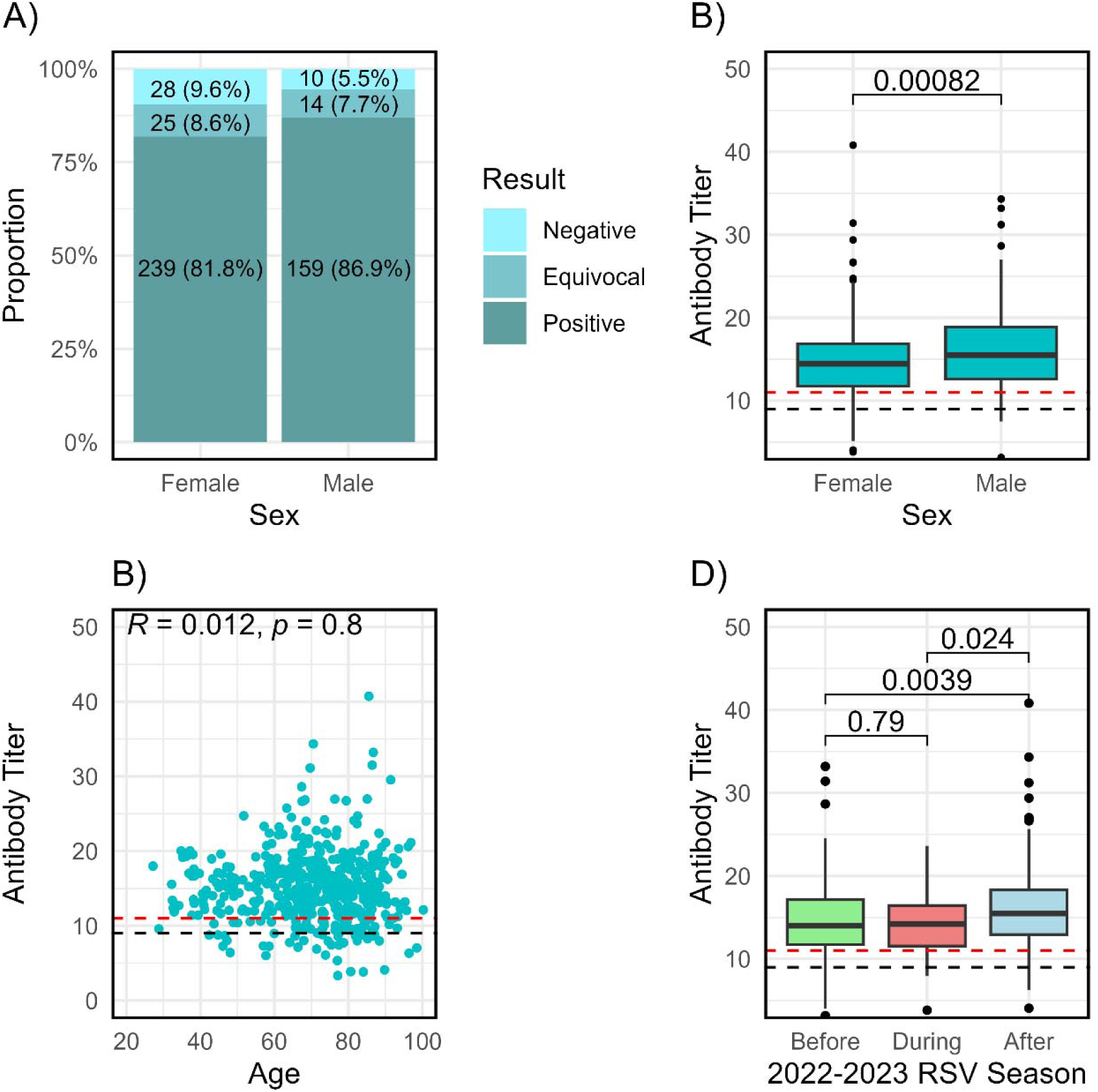
RSV ELISA was conducted on 475 subjects. Males exhibited higher seropositivity rates (A) and significantly elevated mean antibody titers (B) compared to females. No correlation was found between antibody titers and age (C). Antibody titers significantly increased after the 2022-2023 RSV season (D). “Negative” indicates individuals with a confirmed negative result (<9 standard units), while “equivocal” refers to inconclusive results within a zone of uncertainty (9 – 11 standard units), and “positive” represents individuals with a confirmed positive result (> 11 standard units). Dashed lines, when present, represent the threshold values of 9 (black) and 11 (red) standard units.

Results were stratified based on age groups, as summarized in Table 2. Age ranges were divided according to the CDC’s recommendations for RSV vaccination of adults: under 60 years (vaccination recommended for those at increased risk of severe disease and for pregnant women), between 60 and 74 years (single-dose RSV vaccination for those at increased risk of severe disease), and 75 years and above (routine single-dose RSV vaccination for all).[25] Adults age 75+ were the largest subgroup in our cohort (N = 207), with the highest percentage of non-seropositive (negative and equivocal) individuals (18.4%) and the lowest median antibody titer (14.4 SU). Adults under 60 years old were the smallest subgroup in the cohort (N = 108) and had a lower rate of non-seropositivity (14.8%) and higher median antibody titer (15.1 SU). Lastly, adults between 60 and 74 years old (N = 160) demonstrated the lowest non-seropositive antibody result rates (14.4%) and the highest median antibody titer (15.2 SU). None of these differences in seropositivity rate or median Ab titer were statistically significant.

**Table 2.** Seroprevalence Study Results by Age Group. “Negative” indicates individuals with a confirmed negative result (<9 standard units), “Equivocal” refers to inconclusive results within a zone of uncertainty (9 – 11 standard units), and “Positive” represents individuals with a confirmed positive result (> 11 standard units).

| **Age Group** | **Negative** | **Equivocal** | **Positive** | **Sample Size** |
| --- | --- | --- | --- | --- |
| **< 60 years** | 9 (8.3%) | 7 (6.5%) | 92 (85.2%) | 108 |
| **60-74 years** | 11 (6.8%) | 13 (8.0%) | 138 (85.6%) | 160 |
| **75+ years** | 19 (9.2%) | 19 (9.2%) | 169 (81.6%) | 207 |
| **Total** | 38 (8.0%) | 39 (8.2%) | 398 (83.8%) | 475 |

In addition, to examine whether seasonality affects seroprevalence or antibody titers, we evaluated the antibody titers from samples collected before, during, and after the 2022-2023 RSV season in the US Department of Health and Human Services (HHS) Region 5. The RSV season in this Region began the week of October 1st, 2022, and ended in the week of December 3rd, 2022. Mean antibody titers were 14.0 SU pre-season, 14.2 SU during the season, and 15.5 SU post-season (Fig. 1D). Analysis revealed significant differences between median antibody titers from samples collected before and after the 2022-2023 RSV season (14.0 versus 15.5 SU, p = 0.04) and between samples collected during and after that RSV season (p = 0.02). Analysis of seropositivity results and antibody titers per month of sample collection cannot be generalized due to limited sample size and missing data for two months. However, a histogram of qualitative results and antibody titers stratified by month of blood draw are depicted in Supplemental Figure 1.

### 3.3 RSV Antibodies in Previously Diagnosed or Vaccinated Subjects

Figure 2 depicts a comparison of antibody titers across the seroprevalence, diagnosed, and vaccinated cohorts (Fig. 2A), stratified by age groups within each cohort (Figs. 2B-D). The Kruskal-Wallis test comparing mean antibody titers revealed a statistically significant difference between the cohort groups with the vaccinated cohort, demonstrating higher antibody titers than the seroprevalence cohort (16.5 versus 14.8 SU, p < 0.001) and lower antibody titers than the diagnosed cohort (16.5 versus 19.2 SU, p < 0.001). However, no significant differences were observed among age groups within each cohort. Additionally, no differences in antibody titers were found between sexes within the diagnosed or vaccinated cohorts.

**Figure 2.**
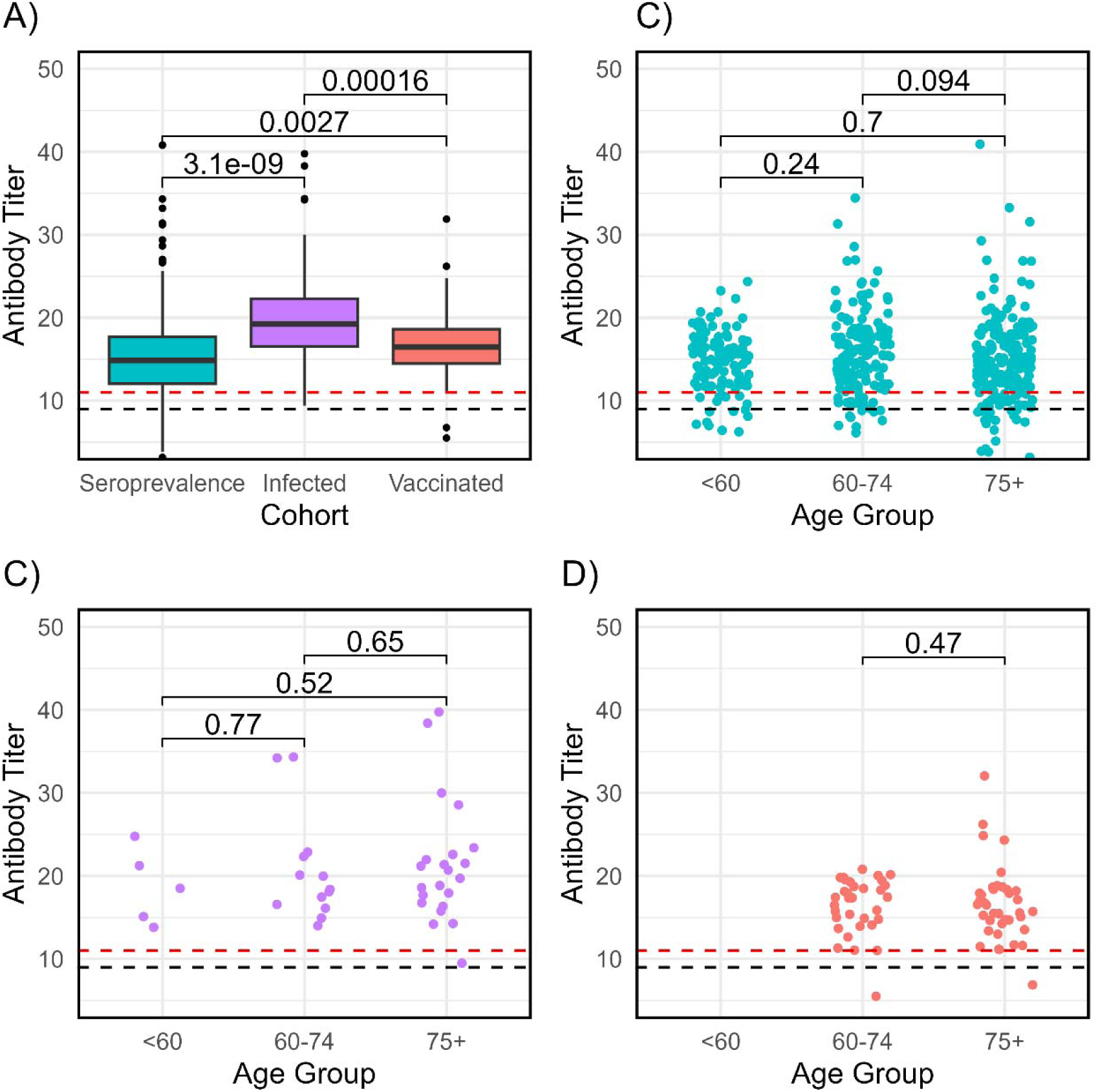
In addition to the seroprevalence cohort (n = 475), cohorts of vaccinated (n = 71) and diagnosed (n = 40) individuals were also studied. Mean antibody titers significantly differed among the seroprevalence, vaccinated, and diagnosed cohorts (A). However, no statistically significant differences in mean antibody titers were observed within the seroprevalence (B), diagnosed (C), or vaccinated (D) cohorts across age groups.

Of the 40 subjects with a previous RSV diagnosis, 39 (97.5%) tested positive, and one (2.5%) had an equivocal result. This subject’s sample was collected over four years after diagnosis, and therefore, the equivocal result may be explained by waning immunity. Of the 71 subjects with a previous RSV vaccination record, 68 (95.8%) tested positive, two (2.8%) tested negative, and one (1.4%) had an equivocal result. The two subjects with negative results had their samples collected 15 or 33 days after the vaccination date, and the subject with an equivocal result had a sample collected 100 days after vaccination. We noted that one of the subjects who tested negative after vaccination had received chemotherapy within 12 months before sample collection. However, neither the other seronegative and the one equivocal subject had any record of an immunocompromising condition. Figure 3 illustrates the relationship between antibody titers and the number of days post-infection (Fig. 3A) or post-vaccination (Fig. 3B). Pearson’s correlation demonstrated that antibody titers significantly decline over time following infection (r = –0.4, p = 0.01). In contrast, there was no observable statistically significant correlation between antibody titers and the number of days post-vaccination (r = 0.14, p = 0.245).

**Figure 3.**
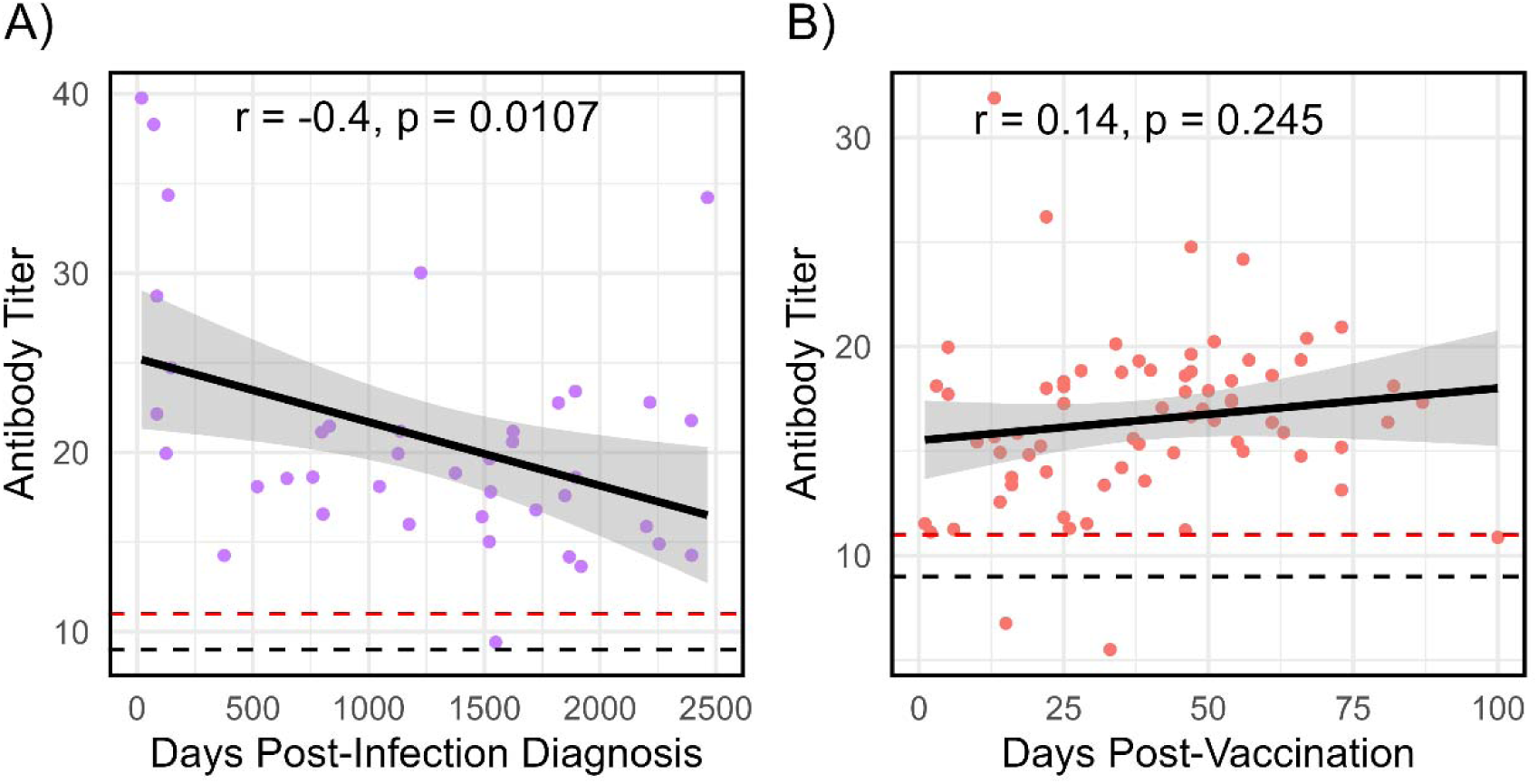
Antibody titers were plotted as a function of days post-infection diagnosis (A) or day post-vaccination (B). A negative correlation was observed in the diagnosed cohort (r = -0.4, p = 0.01), corroborating waning immunity against RSV. No significant relationship was found in the vaccinated cohort.

## 4. DISCUSSION

In this study, we assessed the seroprevalence of RSV IgG antibodies in a cohort of 475 adults with available residual serum samples. The seropositivity rate was lower in the main study (“seroprevalence”) cohort (83.8%) than in the diagnosed or the vaccinated cohorts (97.5% and 95.8%, respectively), indicating that recent exposure to RSV antigen (virus or vaccine) increases the likelihood of a positive qualitative test result for RSV IgG antibodies. The majority of individuals with negative and equivocal results across the three groups did not have records of any potentially immunocompromising condition. Moreover, a comparison between antibody titers of individuals with records of potentially immunocompromising conditions and individuals with no such records yielded similar ranges and mean titers, with no statistical differences between these groups.

Within the vaccinated and infected cohorts, seropositivity rates were not 100%, and only one of the seronegative samples came from an individual with potentially immunocompromising conditions, suggesting that some individuals either did not have a robust humoral response or that rapid waning occurred, and antibody titers dropped below the threshold for positivity in our assay. Figure 3A corroborates RSV waning immunity, demonstrating a negative correlation between antibody titers and the number of days post-infection diagnosis (r = -0.4, p = 0.01). We noted that antibody titers in the vaccinated cohort were only slightly higher than in the seroprevalence cohort, despite recent vaccination. This may be partly due to the whole RSV virus IgG ELISA used in our study. Vaccination will boost pre-fusion F antibody titers only. This expected increase in antibody titer may be masked by our more inclusive assessment of antibody to the whole virus. Antibody titers were not correlated with age in any of the cohorts studied. Additionally, our study revealed a significant increase in antibody titers post-RSV season within the seroprevalence cohort, suggesting the occurrence of undiagnosed RSV infections. This may have resulted from mild or subclinical symptoms that did not necessitate medical attention or from instances where individuals sought care but were not tested for RSV. These results suggest that RSV cases are likely underreported, and the annual burden of RSV disease is almost certainly underestimated. [26, 27]

The strengths of our study are the opportunity to compare the general population to groups of people with known exposure or recent vaccination and its exploration of antibody differences between male and female adults. While this topic has been extensively studied in infants, where male infants are known to be more susceptible to RSV infection and experience more severe disease compared to females, sex differences in adult cohorts have received far less attention.[28] Adult females generally exhibit stronger immune responses and higher antibody titers against viral respiratory infections/vaccinations, such as Influenza and COVID-19.[29, 30] However, our study found that males had higher antibody titers against RSV (15.5 versus 14.5 SU, p < 0.001). Without data on potential differences in exposure risk or time since the last infection within this cohort, it is unclear whether the higher antibody titers observed in males reflect a more robust immune response or are influenced by other factors.

The limitations of our study include the regionality of the cohort (composed only of people who had a blood draw in southeastern Minnesota, which does not represent the US population) and the low representation of minority subjects. Samples were collected in 2022 and 2023 during the COVID-19 pandemic. It is possible that masking, social distancing, and other activities may have changed RSV circulation patterns, affected exposure risk, and altered rates of antibody waning. While CDC RSV surveillance data indicate a significant decrease of RSV (positive tests and hospitalizations) in 2021, RSV incidence rates for 2022-2024 are approximately twice pre-pandemic seasons. These factors should also be considered when comparing our results with other published studies. In addition, the number of days post-infection or vaccination was variable in these convenience cohorts, and baseline antibody levels were not assessed. This, along with the short timeframe between vaccination and sample collection within the vaccinated cohort (<100 days), restricted our ability to draw conclusions about antibody titer trends following vaccination. While the seroprevalence study covered a two-year range and provides important new information on seroprevalence in an older US adult population, there are limitations in the sampling, including the low number studied per month (precluding anything more than an exploratory examination of antibody titer differences across the two RSV seasons) and a potential bias toward less healthy adults as samples were obtained during a medical visit (which could have been routine or not). The sample size for each month of collection was limited, which hindered our ability to detect potential monthly variations in antibody levels. Further investigation in a larger cohort is needed to reliably investigate the seasonal dynamics of antibody titers. Given the difficulty finding a humoral correlate of protection, studies conducting a more comprehensive immune assessment (including T cell responses) in the periphery and mucosal sites are warranted.

On the other hand, this study assessed a contemporary US cohort and is the first to report RSV seroprevalence among the general population in the US, without selecting patients based on respiratory symptoms. Unlike other studies that have focused on screening individuals with symptoms of acute respiratory illness, our study included all patients undergoing a blood draw, regardless of the reason for their visit. Moreover, we could examine differences between sex and age groups, and between the general population from previously diagnosed or vaccinated cohorts, as well as demonstrate that RSV IgG titers significantly increase in the period after the RSV season.

## 5. CONCLUSION

Our study is the first to investigate gender-based differences in RSV antibody titers among adult individuals, with findings indicating that males exhibited higher titers. However, the underlying cause of this observed difference remains undetermined. Our study also demonstrates a significant increase in antibody titers following the RSV season, providing evidence of recent unrecognized/undiagnosed infection. At the same time, over 16.2% of older adults do not have detectable levels of RSV IgG antibodies with this assay, suggesting that they may be more susceptible to infection. Additionally, we demonstrated that there is a negative correlation between antibody titer and the number of days since the last documented RSV infection. While we cannot rule out subsequent exposure, infection, or subclinical disease, these data can be used to estimate rates of waning in the general population. Because infection may occur even when RSV antibodies are detected, and the antibody titer required to prevent illness is currently unknown, it is difficult to accurately estimate the number of older adults vulnerable to RSV infection, even with larger sample sizes. A longitudinal surveillance study with regular antibody measurements and PCR testing to identify infected individuals in a large well-powered cohort could establish a correlate of protection, filling a major knowledge gap, and should be a top research priority. Such a breakthrough would allow us to estimate RSV infection susceptibility more accurately and, therefore, better aid public health authorities in decision-making processes concerning RSV infection prevention and vaccine deployment.

## ACKNOWLEDGMENTS

We would like to thank Dr. Huy Quach for his assistance with data analysis, and Dr. Tamar Ratishvili for helpful comments on an earlier version of this manuscript.

## DATA AVAILABILITY

Data available upon request.

## ETHICAL APPROVAL

The described procedures were approved by the Mayo Clinic Institutional Review Board (IRB# 08-007049).

## CONTRIBUTIONS

**Lara I. Teodoro:** Writing - original draft. Investigation, Formal analysis, Data curation, Visualization. Conceptualization. **Inna G. Ovsyannikova:** Writing - review & editing. **Diane E. Grill:** Writing - review & editing. Data curation. **Gregory A. Poland:** Writing - review & editing, Conceptualization. **Richard B. Kennedy:** Writing - review & editing, Supervision. Conceptualization. Funding.

## SUPPLEMENTAL

**Supplemental Figure 1.**
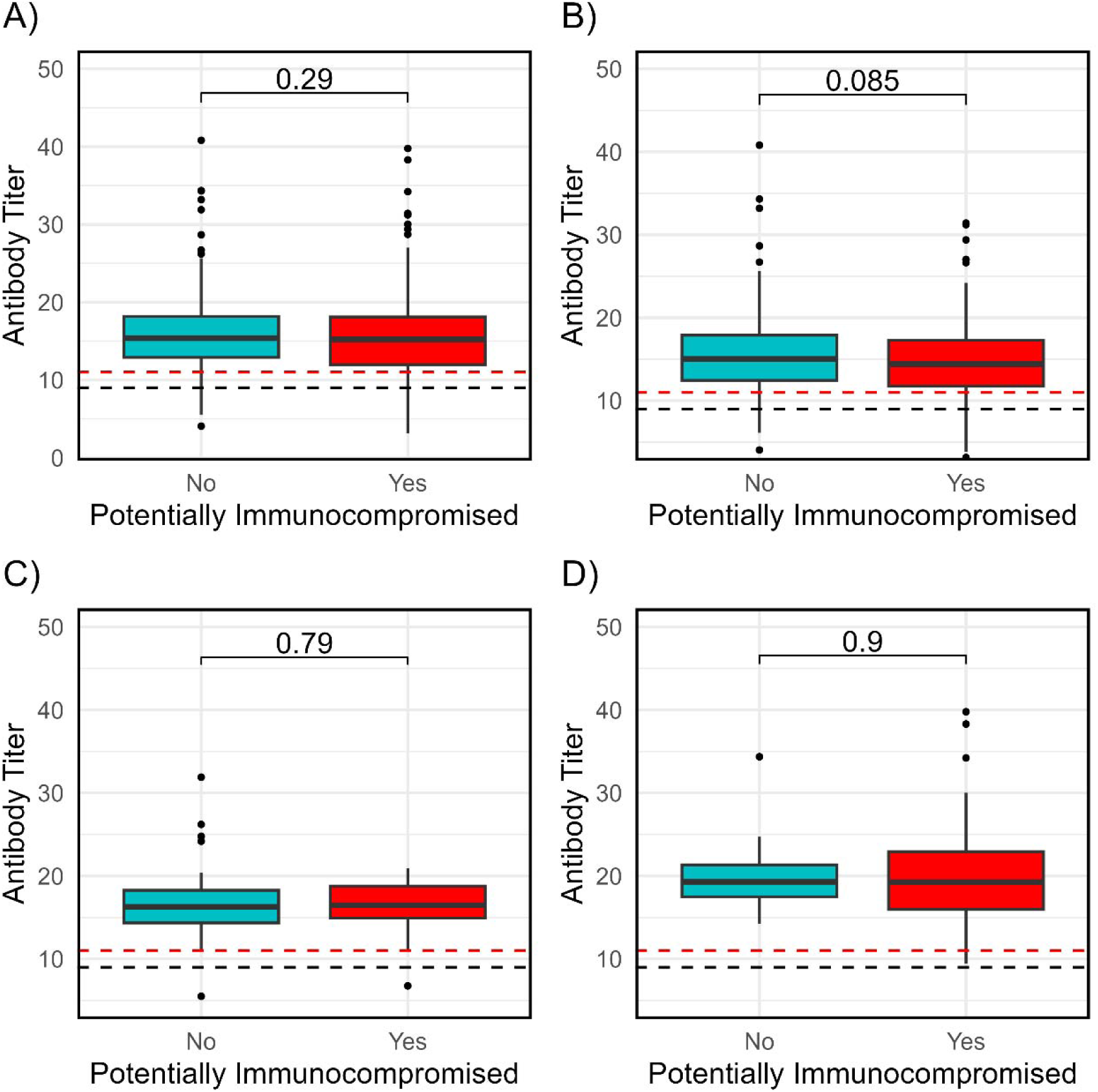
Comparison of antibody titers between individuals with records of potentially immunocompromising conditions and those without such records revealed similar ranges, with no significant differences observed overall (A), or within the seroprevalence (B), vaccinated (C), or infected (D) cohorts.

**Supplemental Figure 2.**
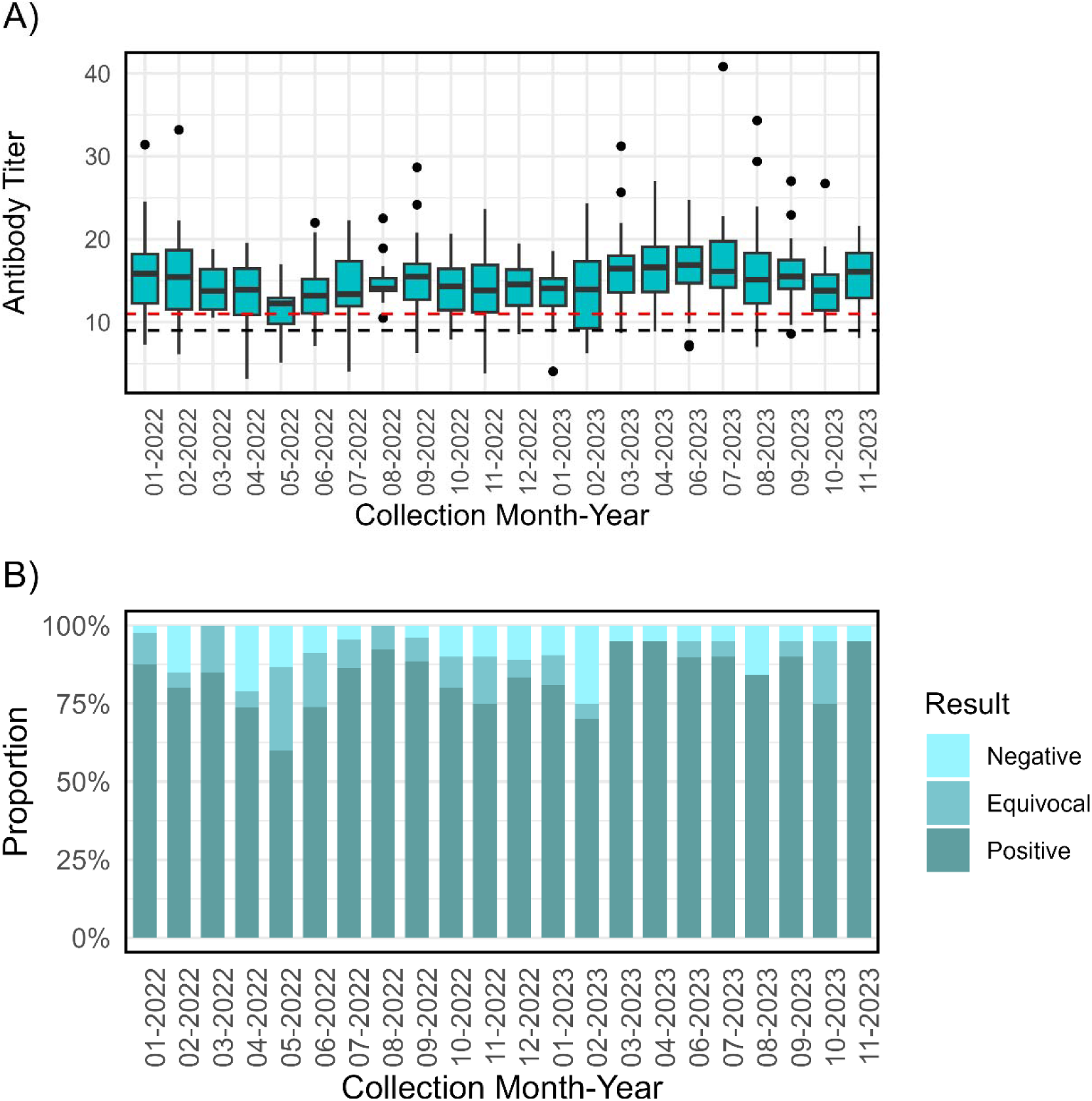
Histogram of antibody titers (A) and qualitative results (B) stratified by month of blood draw.

## REFERENCES

[1] Rima, B., P. Collins, A. Easton, R. Fouchier, G. Kurath, R.A. Lamb, et al. ICTV Virus Taxonomy Profile: Pneumoviridae. J Gen Virol, 2017. 98(12): p. 2912–2913.

[2] Mufson, M.A., C. Orvell, B. Rafnar, and E. Norrby. Two distinct subtypes of human respiratory syncytial virus. J Gen Virol, 1985. 66 (Pt 10): p. 2111–24.

[3] National Institute of Allergy and Infectious Diseases. Respiratory Syncytial Virus (RSV). 2022 July 22, 2022 November 1, 2024]; Available from: https://www.niaid.nih.gov/diseases-conditions/respiratory-syncytial-virus-rsv.

[4] Rosas-Salazar, C., T. Chirkova, T. Gebretsadik, J.D. Chappell, R.S. Peebles, Jr., W.D. Dupont, et al. Respiratory syncytial virus infection during infancy and asthma during childhood in the USA (INSPIRE): a population-based, prospective birth cohort study. Lancet, 2023. 401(10389): p. 1669–1680.

[5] Centers for Disease Control and Prevention. Respiratory Syncytial Virus Infection (RSV). 2024 August 30, 2024 November 1, 2024]; Available from: https://www.cdc.gov/rsv/about/index.html.

[6] Falsey, A.R. and E.E. Walsh. Respiratory syncytial virus infection in elderly adults. Drugs Aging, 2005. 22(7): p. 577–87.

[7] Hall, C.B., E.E. Walsh, C.E. Long, and K.C. Schnabel. Immunity to and frequency of reinfection with respiratory syncytial virus. J Infect Dis, 1991. 163(4): p. 693–8.

[8] Piedra, P.A., A.M. Jewell, S.G. Cron, R.L. Atmar, and W.P. Glezen. Correlates of immunity to respiratory syncytial virus (RSV) associated-hospitalization: establishment of minimum protective threshold levels of serum neutralizing antibodies. Vaccine, 2003. 21(24): p. 3479–82.

[9] Centers for Disease Control and Prevention. Healthcare Providers: RSV Vaccination for Adults 60 Years of Age and Over. 2024 July 3, 2024 November 1, 2024]; Available from: https://www.cdc.gov/vaccines/vpd/rsv/hcp/older-adults.html.

[10] Food and Drug Administration. AREXVY. Vaccines, Blood & Biologics August 22, 2024 November 1, 2024]; Available from: https://www.fda.gov/vaccines-blood-biologics/arexvy.

[11] Food and Drug Administration. ABRYSVO. Vaccines, Blood & Biologics October 23, 2024 November 1, 2024]; Available from: https://www.fda.gov/vaccines-blood-biologics/abrysvo.

[12] Food and Drug Administration. MRESVIA. Vaccines, Blood & Biologics October 4, 2024 November 1, 2024]; Available from: https://www.fda.gov/vaccines-blood-biologics/vaccines/mresvia.

[13] Widmer, K., M.R. Griffin, Y. Zhu, J.V. Williams, and H.K. Talbot. Respiratory syncytial virus- and human metapneumovirus-associated emergency department and hospital burden in adults. Influenza Other Respir Viruses, 2014. 8(3): p. 347–52.

[14] Widmer, K., Y. Zhu, J.V. Williams, M.R. Griffin, K.M. Edwards, and H.K. Talbot. Rates of hospitalizations for respiratory syncytial virus, human metapneumovirus, and influenza virus in older adults. J Infect Dis, 2012. 206(1): p. 56–62.

[15] Tong, S., C. Amand, A. Kieffer, and M.H. Kyaw. Incidence of respiratory syncytial virus related health care utilization in the United States. J Glob Health, 2020. 10(2): p. 020422.

[16] Sieling, W.D., C.R. Goldman, M. Oberhardt, M. Phillips, L. Finelli, and L. Saiman. Comparative incidence and burden of respiratory viruses associated with hospitalization in adults in New York City. Influenza Other Respir Viruses, 2021. 15(5): p. 670–677.

[17] Nolen, L.D., S. Seeman, C. Desnoyers, C. DeByle, J. Klejka, D. Bruden, et al. Respiratory syncytial virus and influenza hospitalizations in Alaska native adults. J Clin Virol, 2020. 127: p. 104347.

[18] Mesa-Frias, M., C. Rossi, B. Emond, B. Bookhart, D. Anderson, S. Drummond, et al. Incidence and economic burden of respiratory syncytial virus among adults in the United States: A retrospective analysis using 2 insurance claims databases. J Manag Care Spec Pharm, 2022. 28(7): p. 753–765.

[19] McClure, D.L., B.A. Kieke, M.E. Sundaram, M.D. Simpson, J.K. Meece, F. Sifakis, et al. Seasonal incidence of medically attended respiratory syncytial virus infection in a community cohort of adults >/=50 years old. PLoS One, 2014. 9(7): p. e102586.

[20] Jackson, M.L., E. Scott, J. Kuypers, A.K. Nalla, P. Roychoudury, and H.Y. Chu. Epidemiology of Respiratory Syncytial Virus Across Five Influenza Seasons Among Adults and Children One Year of Age and Older-Washington State, 2011/2012-2015/2016. J Infect Dis, 2021. 223(1): p. 147-156.

[21] Branche, A.R., L. Saiman, E.E. Walsh, A.R. Falsey, W.D. Sieling, W. Greendyke, et al. Incidence of Respiratory Syncytial Virus Infection Among Hospitalized Adults, 2017-2020. Clin Infect Dis, 2022. 74(6): p. 1004-1011.

[22] US Department of Health and Human Services. Regional Offices. December 9, 2024 December 9, 2024]; Available from: https://www.hhs.gov/about/agencies/ogc/offices/regional-offices/index.html.

[23] Centers for Disease Control and Prevention. Interactive Dashboard. The National Respiratory and Enteric Virus Surveillance System (NREVSS) December 6, 2024 December 9, 2024]; Available from: https://www.cdc.gov/nrevss/php/dashboard/index.html.

[24] Midgley, C.M., A.K. Haynes, J.L. Baumgardner, C. Chommanard, S.W. Demas, M.M. Prill, et al. Determining the Seasonality of Respiratory Syncytial Virus in the United States: The Impact of Increased Molecular Testing. J Infect Dis, 2017. 216(3): p. 345–355.

[25] Centers for Disease Control and Prevention. RSV Vaccines. August 30, 2024 [cited November 1, 2024; Available from: https://www.cdc.gov/rsv/vaccines/index.html.

[26] Havers, F.P., M. Whitaker, M. Melgar, H. Pham, S.J. Chai, E. Austin, et al. Burden of Respiratory Syncytial Virus-Associated Hospitalizations in US Adults, October 2016 to September 2023. JAMA Netw Open, 2024. 7(11): p. e2444756.

[27] Carrico, J., K.A. Hicks, E. Wilson, C.A. Panozzo, and P. Ghaswalla. The Annual Economic Burden of Respiratory Syncytial Virus in Adults in the United States. J Infect Dis, 2024. 230(2): p. e342–e352.

[28] Ursin, R.L. and S.L. Klein. Sex Differences in Respiratory Viral Pathogenesis and Treatments. Annu Rev Virol, 2021. 8(1): p. 393–414.

[29] Voigt, E.A., I.G. Ovsyannikova, R.B. Kennedy, D.E. Grill, K.M. Goergen, D.J. Schaid, et al. Sex Differences in Older Adults’ Immune Responses to Seasonal Influenza Vaccination. Front Immunol, 2019. 10: p. 180.

[30] Qi, S., C. Ngwa, D.A. Morales Scheihing, A. Al Mamun, H.W. Ahnstedt, C.E. Finger, et al. Sex differences in the immune response to acute COVID-19 respiratory tract infection. Biol Sex Differ, 2021. 12(1): p. 66.

