## Supplemental Data 1 for "Seroprevalence of RSV Antibodies in a Contemporary (2022 – 2023) Cohort of Adults"

**SUPPLEMENTAL FIGURES**

**Supplemental Figure 1.** Comparison of antibody titers between individuals with records of potentially immunocompromising conditions and those without such records revealed similar ranges, with no significant differences observed overall (A), or within the seroprevalence (B), vaccinated (C), or infected (D) cohorts.

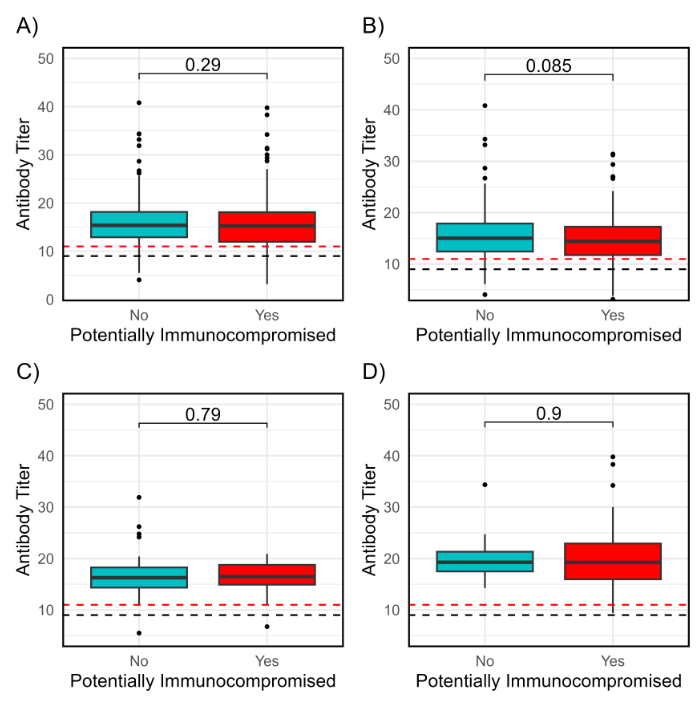

**Supplemental Figure 2.** Histogram of antibody titers (A) and qualitative results (B) stratified by month of blood draw.

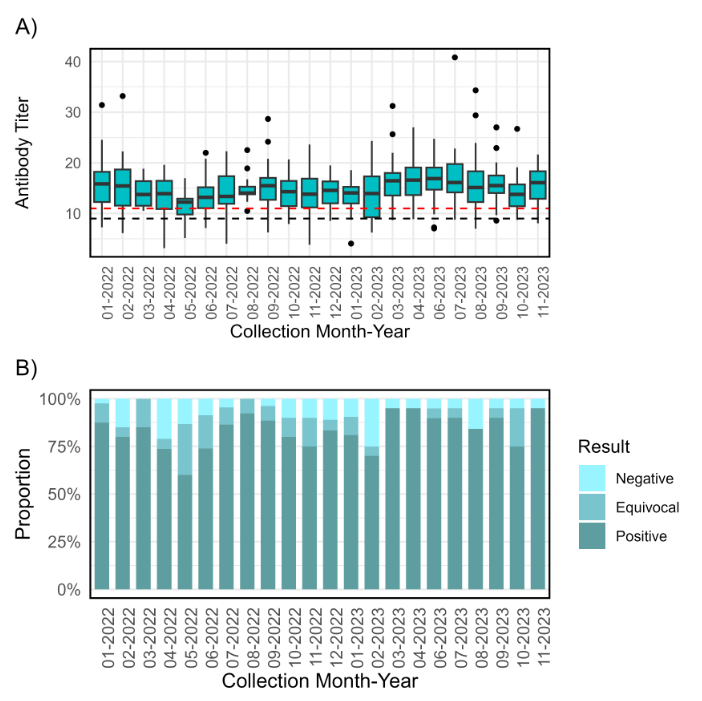
